# Association of quantified cardiovascular health with all-cause mortality in prediabetic patients

**DOI:** 10.1101/2024.07.31.24311259

**Authors:** Aomiao Chen, Qiuyu He, Yichuan Wu, Jiaqi Chen, Xiaoqin Ma, Lingyuan Hu, Geningyue Wang, Zhuotong Wang, Zongji Zheng, Yijie Jia

## Abstract

**Aim:** We aimed to explore the association between all-cause mortality and cardiovascular health (CVH) lifestyle interventions (as accurately quantified by Life’s essential 8) in prediabetic patients and to observe the dose-response relationship of the potential association.

**Methods and Participants:** The retrospective study included 5344 participants with prediabetes (age: 52.9 ±15.8 years; (51.6% of men)). The Life’s essential 8 (LE8) score includes four health indicators and four health behaviors. We calculated Cox proportional hazard ratios (HRS) for all-cause mortality in subgroups of high CVH (≥80), low CVH (≤50), and moderate CVH (50-79) based on the CVH quantification score of LE8, and explored the dose-response relationship of potential associations. We also performed separate analyses of the associations of all-cause mortality with each LE8 components and CVH health behaviors and indicators.

**Results:** In the median follow-up period of 8.33 years, 658 deaths occurred. Compared with participants with high CVH, the covariate-adjusted HR(95%CI) for participants with moderate and low CVH were 2.55(1.23-5.31) and 3.92 (1.70-9.02), respectively. There was a dose-response relationship between the improvement of CVH status and the reduction of all-cause mortality (P-overall < 0.0001, P-nonliner = 0.7989). The improvement of CVH health behaviors has a more significant protective effect on patients with prediabetes than CVH health indicators.

**Conclusion:** High CVH status, quantified by LE8, has a significant effect on preventing mortality outcomes in prediabetic adults in the U.S.

## **I.** Background

Prediabetes is defined as a glucose condition which is a high-risk metabolic state of diabetes^1^. It is estimated that by the year of 2030, 470 million people will suffer from prediabetes in the world^2, 3^, and more than 70% of prediabetic patients will eventually develop diabetes^4^. Moreover, prior guidelines and two meta-analyses involving more than one million people have shown that patients with prediabetes indeed encounter elevated risk for cardiovascular disease (CVD)^2, 5, 6^.

Previous guidelines and numerous large cohort studies have focused on identifying effective health interventions for diabetes, which are essential to reduce the medical burden and improve survival outcomes^7–10^. However, there is heterogeneity in the health status of individual patients’ lifestyle that cannot be ignored^11^. Using an intervention-control design based on a single cutoff point, studies to determine the effectiveness of health interventions without accurate quantification of the health assessment process are likely to produce diametrically opposite results^8, 12^. This is most likely due to differences between the two studies in the setting of cut-off points for health assessment (For example, the physical activity requirement for healthy young adults is an additional 280 minutes of moderate-intensity activity per week in the DQDPOS and 150 minutes per week in the DPPOS)^9, 13^. Therefore, we believe that quantified healthy living status based on uniform standards will be useful to confirm whether health interventions can preventatively reduce the risk of death in prediabetic patients. To further improve the global cardiovascular health (CVH), the AHA recently recommended the Life’s Essential 8 (LE8) score to quantify cardiovascular health, which includes four CVH behaviors (diet, sleep, exercise, smoking cessation) and four ideal CVH indicators (blood lipids, blood glucose, blood pressure, body mass index)^14, 15^. We believe that LE8 can achieve accurate quantification of CVH, which is beneficial to clarify whether health interventions in prediabetic patients can reduce the overall risk of death, and its intuitive score is important for urging the prediabetic population to achieve and maintain optimal CVH status^16^.

Based on its efficacy in health assessment, LE8 has been widely used in prevention and prognosis related studies^17–20^. However, to our knowledge, there are few study has investigated the association between LE8 and all-cause mortality in patients with T2DM^21^. We conclude that CVH intervention should be strengthened as early as possible in not only T2DM patients, but also pre-diabetic patients who are at risk of death and CVD. Especially in prediabetic patients, early health intervention can often achieve better results^4^. However, in medical practice, many prediabetic patients with CVD will be forced to change their lifestyle due to functional limitations or psychological reasons^22–24^. These patients will be more difficult to achieve a high CVH state (LE8≥80) or even a moderate CVH state (LE8≥50) as defined by the AHA. In addition, because special factors are not considered by LE8 such as the obesity paradox may exist in the disease patient population^25^, it is important to analyze the relationship between the individual components of LE8 and mortality to clarify their efficacy. Finally, CVH behavioral intervention itself can improve CVH indicators and is more accessible in the components of LE8^26^. However, assessment of this component has not been implemented in prediabetic population.

Therefore, we studied a nationally representative queue, in order to realize the following three goals. First, we aimed to determine whether quantified high CVH status, based on LE8, could preventatively reduce the risk of all-cause mortality in prediabetic patients. Second, the dose-response form of this preventive effect will be confirmed, which will benefit prediabetic patients who can only achieve limited CVH improvement. Finally, we aimed to explore the associations of health behaviors, health indicators and individual components of LE8 with mortality risk in order to identify the most effective intervention strategies.

## **II.** Research Design and Methods

### 1. Study Design and Population

The data and guidelines used in this analysis are freely available from NCHS:https://www.cdc.gov/nchs/nhanes/index.htm. The National Health and Nutrition Examination Survey (NHANES) is a nationally representative study which was designed to investigate the nutritional status and health in the United States. Research design and methods has been described in detail (http://www.cdc.gov/nchs/nhanes/about_nhanes.htm.). The NHANES study protocol has been approved by the National Center for Health Statistics Institutional Review board, and each participant provided formal informed consent.

Five NHANES cycles from 2007 to 2016 with 29201 adult participants was uesd as baseline. After excluding participants with insufficient data on diabete-related biochemical markers(n = 16295), serum creatinine (n = 160), cardiovascular healthy lifestyle (n = 2333), and mortality follow-up (n = 5), weight data missing (n = 317), and 10091 participants were included in the analysis.

According to ADA’s criteria and previous studies^14, 27, 28^, prediabetes was defined as a self-reported physcian-diagnosed prediabetic state or an HbA1c level form 5.7% to 6.4% or a fasting blood glucose level form 100mg/dL to 125mg/dL. Since this study targeted a prediabetic population, 5344 participants were included in this study after screening.

### 2. Assessment of cardiovascular health

Cardiovascular health (CVH) was assessed using the LE8 score^22^, which consists of: Physical activity, Diet health, nicootine exposure, BMI, Sleep health, Blood glucose, Blood lipid, Blood pressure. LE8 scores were calculated as the average of the eight component scores, ranging from 100 to 0. According to recommendations from AHA, CVH status was classified according to the LE8 score: 80-100 as high CVH, 0-49 as low CVH, and 50-79 as moderate CVH.

Eight components were scored according to different metrics (Supplementary Table 1). The Healthy Eating Index 2020 (HEI-2020) was used to calculate the diet score^29^, and dietary information was obtained by self-reported food frequency questionnaire. Information on self-reported physical activity, nicotine exposure, and sleep duration was collected by NHANES questionnaire. Height, weight, and blood pressure were measured at the examination centers using standard instruments. Non-HDL cholesterol was used to calculate Lipid score. Blood glucose was scored by means of fasting plasma glucose or HbA1c measured by standard methods.

### 3. Covariate assessment

Covariates included demographic variables, health-related exposures, and underlying diseases. Demographic variables include years of age, gender, ethnicity, income and poverty rate. Health related exposure include drinking, smoking, BMI(kg/m2), total cholesterol. Basic diseases including diabetes, hypertension, cardiovascular disease (myocardial infarction, congestive heart failure, angina attack, or coronary heart disease, as diagnosed by a physician), and anemia.

### 4. Mortality assessment

Baseline data from the NHANES from 2007 through 2016 were linked to the National Death Index (NDI) through December 31, 2019, to determine survival.

### 5. Statistical analysis

All analyses accounted for the complex sampling design of the NHANES and used fasting subsample weights(wtsaf10yr) to ensure nationally representative results. CVH status was classified according to the LE8: 80-100 as high CVH, 0-49 as low CVH, and 50-79 as moderate CVH status^15^.

First, Kaplan-Meier method was used to visually analyze the cumulative mortality of prediabetic patients with different CVH status categories. We then developed multivariable COX proportional hazards models to assess the association bwtween CVH status and all-cause mortality based on follow-up time. The model passed Schoenfeld residual tests for statistical compliance (Supplementary Figure 1), and C statistics were calculated. We included demographic variables, health-related exposures, and basic diseases as covariates to adjust for confounding factors.

Next, to analyze the dose-response relationship between all-cause mortality and the total CVH score LE8 in prediabetic patients, restricted cubic spline models (RCS) of 3 sections (90th, 50th, and 10th percentiles) were applied. Models were evaluated according to Akaike information criterion (AIC) and tested for nonlinearity. To test the generalizability and sensitivity of this relationship, we repeated the RCS models, stratified according to age, sex, income and poverty ratio, and the presence or absence of cardiovascular disease or anemia. The second sensitivity analysis eliminates the survival time less than one year follow-up of adults, to evaluate whether the results under the influence of reverse causation. In the third sensitivity analysis, all participants with CVD were excluded, because the disease itself may force patients to make lifestyle changes, such as physical activity restrictions.

Finally, to investigate the best protective factors for patients with prediabetes, and to analyze the protective effect of health behaviors and health indicators, each LE8 component score (grouped by whether it was 50 or higher) and the mean score on health behaviors and on health indicators were included as additional covariates in the COX proportional-hazards model. R software was used to perform all analyses (Version: R 4.2.1; https://www.r-project.org/). Double-sided P-values of less than 0.05 were considered to indicate statistical significance.

## **III.** Results

### 1. Baseline characteristics

Table 1 describes the baseline characteristics of the total cohort of participants. Only 7.0% of participants were classified as having high CVH. Prediabetic adults with better CVH status were more likely to be male, white people with higher family income level, do not drink alcohol or smoke, better control of blood lipids, blood glucose, and blood pressure, and do not have basic diseases.

**Table 1.**
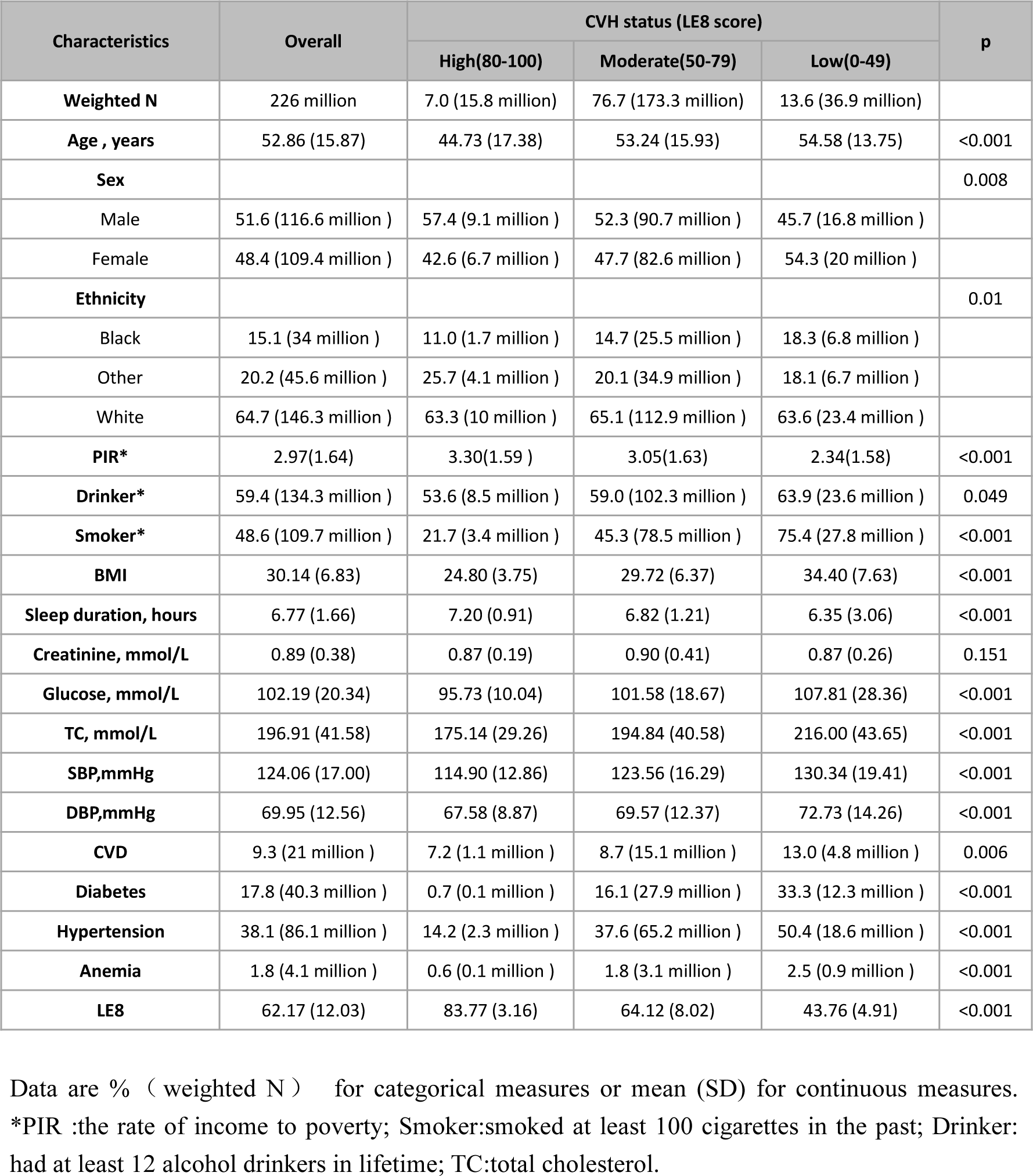
Baseline characteristics of the weighted study population classified by CVH status.

### 2. Association between CVH status and mortality in patients with prediabetes

During a median follow-up period of 8.33 years (95%CI, 8.25-8.50), 658 deaths occurred. As shown in Figure1, Kaplan-Meier survival curves differed by CVH status (P<0.001, Log-rank). The survival curves of the high CVH group were significantly higher than those of the moderate and low CVH groups.

**Figure 1.**
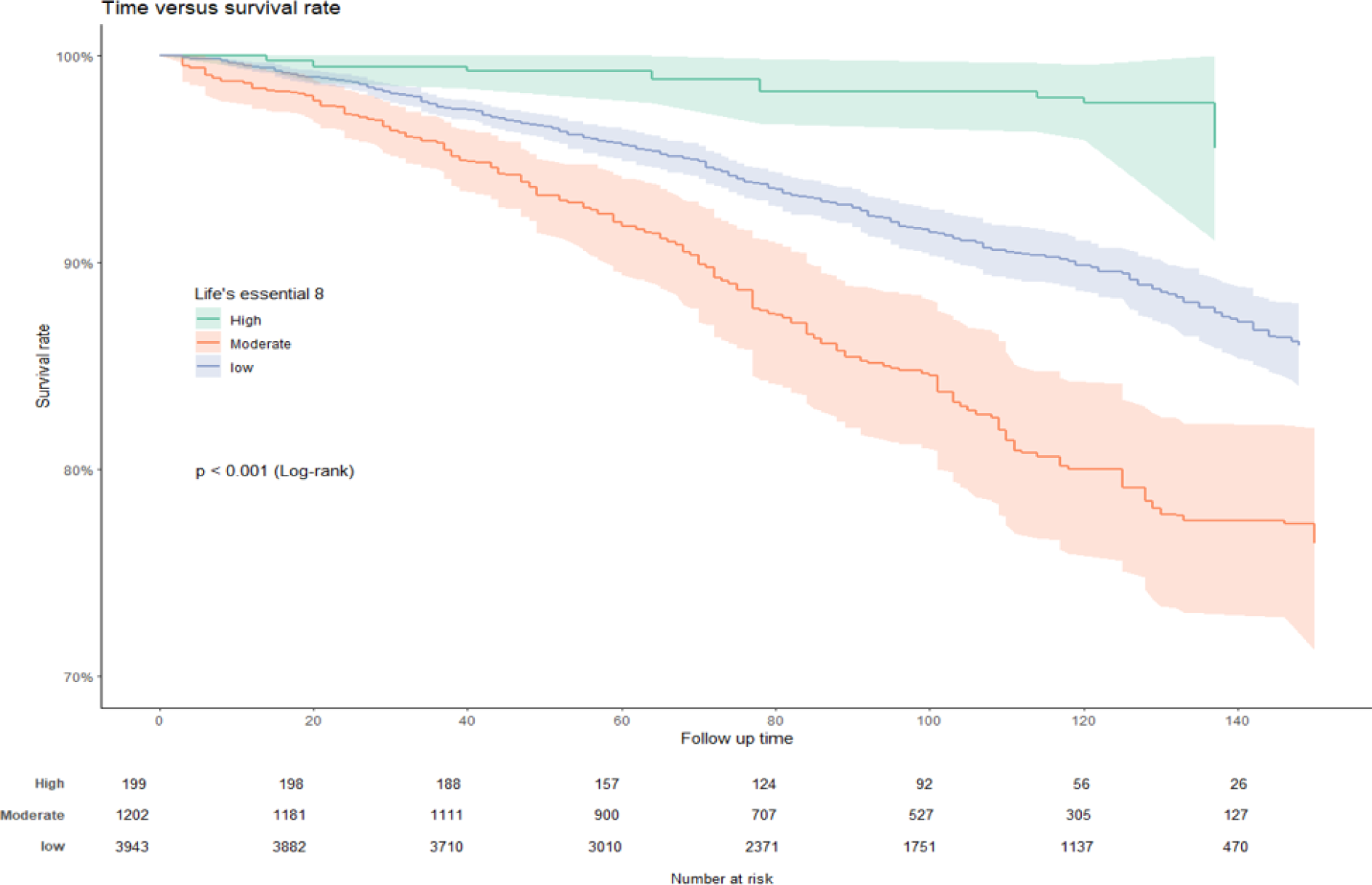
Association of different CVH status and cumulative mortality in prediabetic patients.

### 3. Association between CVH status and risk of mortality in patients with prediabetes

We developed multivariable COX proportional hazards models, as shown in Table 2, in which worsening CVH status leading to an increased risk of death. After adjusting for all potential covariates (Model 3, C-statistic: 0.8252166), the risk of death in the moderate CVH group and the low CVH group was respectively 2.55 times (HR: 2.55, 95%CI: 1.23-5.31) and 3.92 times (HR:3.92, 95%CI:1.70-9.02) higher than that in the high CVH group.

**Table 2.**
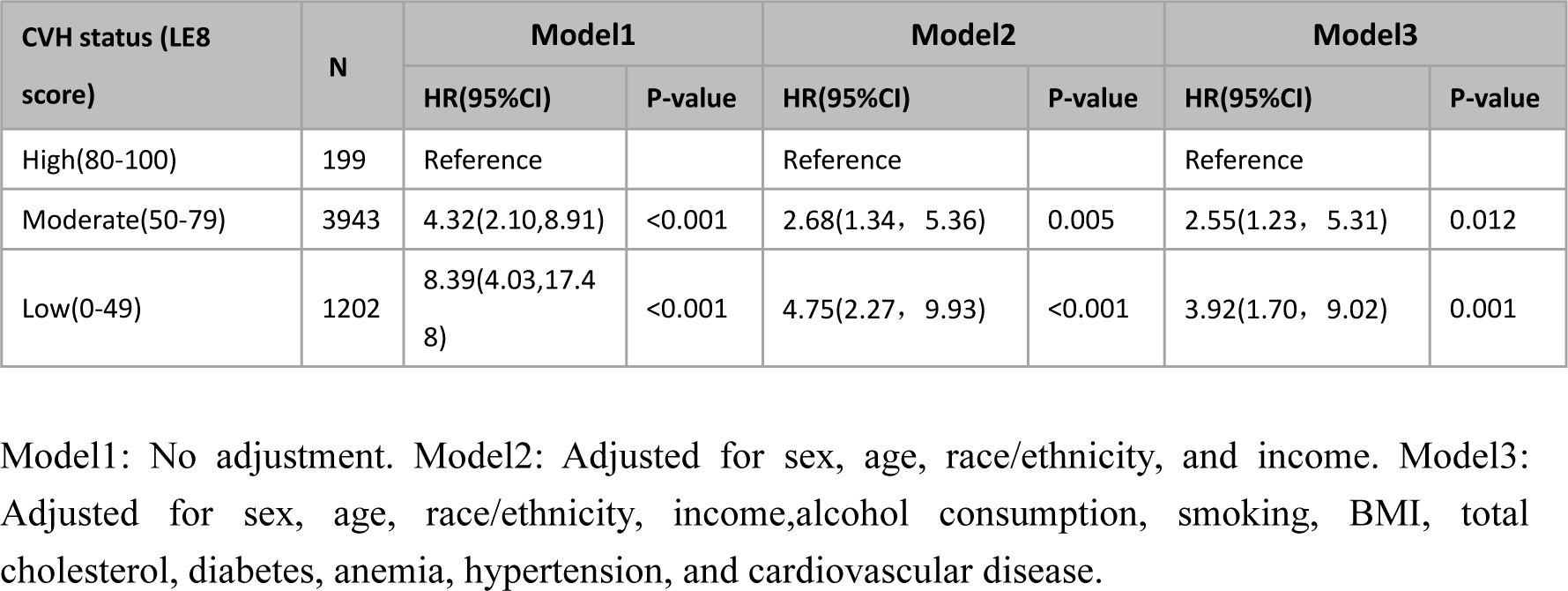
Hazard ratios of cardiovascular health status to all-cause mortality in prediabetes.

| CVH status (LE8 score) | N | Model1 |  | Model2 |  | Model3 |  |
| --- | --- | --- | --- | --- | --- | --- | --- |
|  |  | HR(95%CI) | P-value | HR(95%CI) | P-value | HR(95%CI) | P-value |
| High(80-100) | 199 | Reference |  | Reference |  | Reference |  |
| Moderate(50-79) | 3943 | 4.32(2.10,8.91) | <0.001 | 2.68(1.34, 5.36) | 0.005 | 2.55(1.23, 5.31) | 0.012 |
| Low(0-49) | 1202 | 8.39(4.03,17.48) | <0.001 | 4.75(2.27, 9.93) | <0.001 | 3.92(1.70, 9.02) | 0.001 |

Model1: No adjustment. Model2: Adjusted for sex, age, race/ethnicity, and income. Model3: Adjusted for sex, age, race/ethnicity, income, alcohol consumption, smoking, BMI, total cholesterol, diabetes, anemia, hypertension, and cardiovascular disease.

### 4. Dose-response relationship between CVH total score (Life’s essential 8) and all-cause mortality in prediabetic patients

RCS modeling and visualization were adopted to investigate the dose-response relationship in prediabetic patients. Multivariable adjusted RCS models showed a significant linear association between LE8 and all-cause mortality (P-overall < 0.0001, P-nonliner = 0.7989, Figure 2a). Akaike information criterion evaluation showed that the model had good statistical power with AIC = 19.00. Through calculation, every 10 points average increment, can avoid the risk for all-cause mortality in patients with up to 30%.

**Figure 2.**
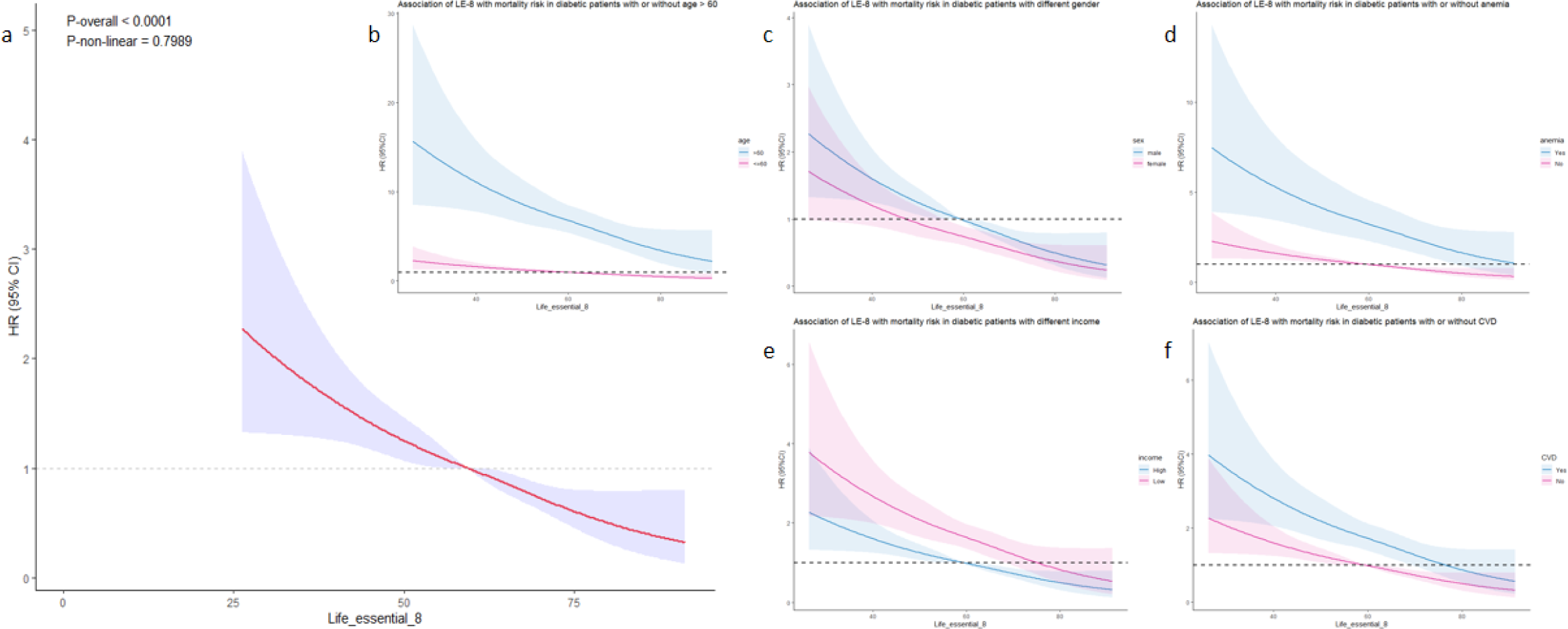
Dose-response relationship between the total CVH score of Life’s essential 8 and all-cause mortality in patients with prediabetes. Panel b-f: Stratified analysis was performed according to age group (greater than 60 years old or not), gender group (male or female), income to poverty rate (higher than the study population median 2.06 or not), and whether there was cardiovascular disease (defined as any one of myocardial infarction, congestive heart failure, angina attack, or coronary heart disease, as diagnosed by a physician) and anemia(defined as hemoglobin < 12mg/dL in men and < 11mg/dL in women).

Stratified analyses showed that the LE8 score was linearly associated with all-cause mortality across all strata (Figure 2b through 2f). In addition, after eliminated all CVD patients, dose-response relationship between LE8 and all-cause mortality is still significant(P-overall < 0.0001, P-nonliner = 0.8749, Supplementary Figure 2a). To avoid the effect of reverse causality, we also examined the association of LE8 with all-cause mortality in the subgroup of patients who had at least 1 year of follow-up, and the results were generally consistent with those in the overall population (P-overall < 0.0001, P-nonliner = 0.7198; Supplementary Figure 2b).

### 5. Associations of individual components of CVH, health behaviors, and health indicators with all-cause mortality in patients with prediabetes

We included individual components of CVH status as covariates in the multivariable COX regression model, and the results are shown in the forest plot (Figure 3a). We found that among health behaviors, higher scores of sleep (HR: 0.76, 95%CI: 0.62-0.92), exercise (HR: 0.49,95%CI: 0.38-0.62) and smoking (HR: 0.73,95%CI: 0.54-0.98) were associated with lower risk of all-cause mortality. However, no significant association was found for Diet Score. In terms of health indicators, we found that prediabetic patients with higher scores were at higher risk of all-cause mortality, such as BMI Score (HR: 1.37,95%CI: 1.13-1.66) and Bodylipid Score (HR: 1.21,95%CI: 1.01-1.44). Moreover, we noticed a protective association in the high Glucose Score (HR: 0.63, 95%CI: 0.49-0.80) but no significant association in the Bloodpressure Score.

**Figure 3.**
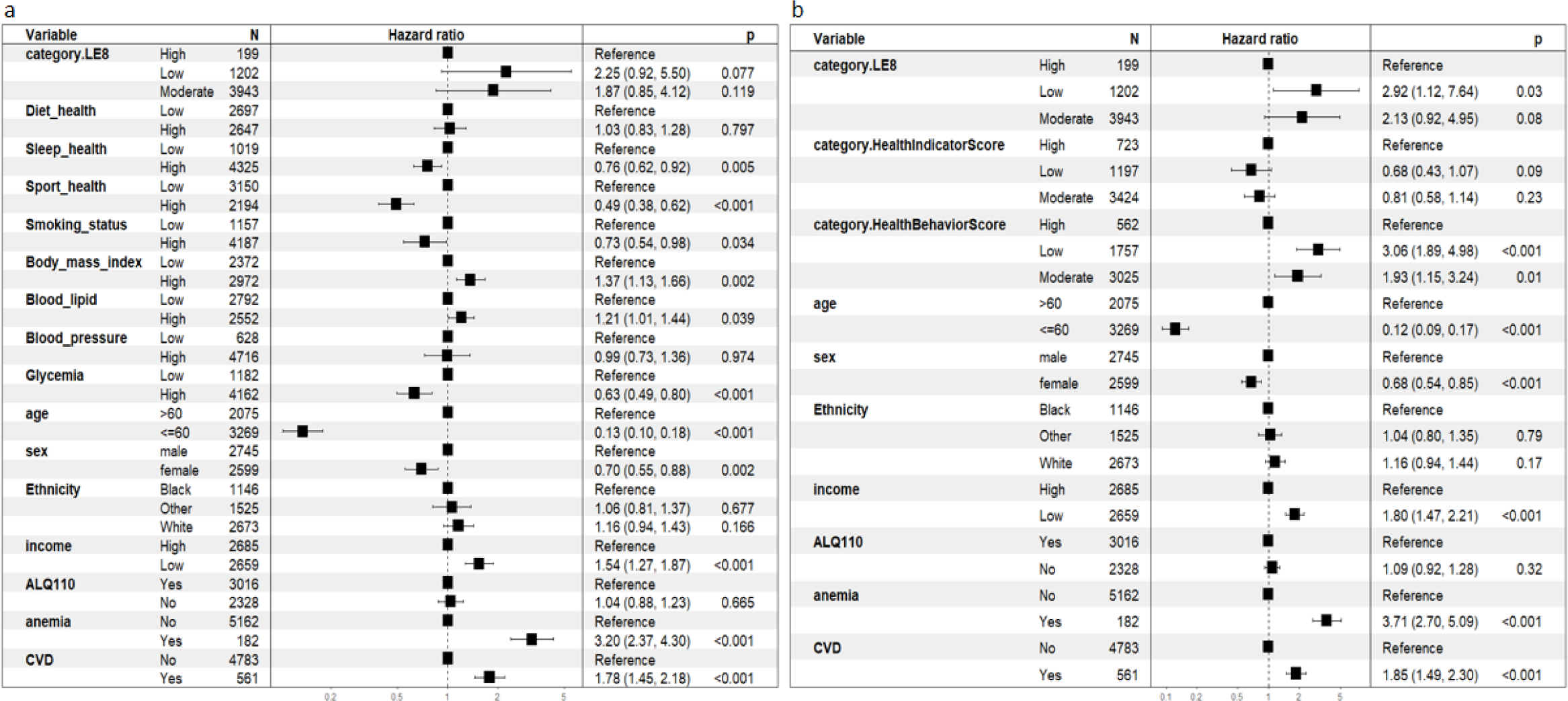
Association of individual CVH component scores, health behaviors or indicators and all-cause mortality in patients with prediabetesa. Panel a-association of individual CVH component scores and all-cause mortality in patients with prediabetes (high for ≥ 50 points, otherwise low). Panel b-association of mean score of health behaviors or indicators and all-cause mortality in patients with prediabetes. Covariates included age (greater than 60 years old or not), gender (male or female), ethnicity (black, white, or other people), income and poverty rate (higher than the study population median 2.06 or not), cardiovascular disease (defined as any one of myocardial infarction, congestive heart failure, angina attack, or coronary heart disease, as diagnosed by a physician), and anemia (defined as hemoglobin < 12mg/dL in men and < 11mg/dL in women). ALQ110: at least 12 cups of alcohol in a life time. CVD: cardiovascular disease.

Because of the aforementioned heterogeneity in the associations between scores on health behaviors and health indicators, we further explored the association between mean scores on health behaviors and health indicators and all cause-mortality by constructing a multivariate COX regression model (Figure 3b). Compared with participants with high average scores of health behavior, participants with medium and low average scores of health behavior had 1.93 times (HR: 1.93, 95%CI: 1.15-3.24) and 3.92 times (HR:3.06, 95%CI:1.89-4.98) risk of death, respectively. However, the protective effect of higher mean scores on health indicators was no longer significant (Figure 3b).

## **IV.** Discussion

Based on a nationally representative sample from the NHANES, we found that high CVH status quantified by LE8, had a significant effect on preventing mortality outcomes in patients with prediabetes. In the linear dose-response relationship, the avoidable mortality risk in prediabetic patients reached 30% for every 10-point LE8 increment on average. Improving CVH health behaviors has a more significant protective effect on prediabetic patients than CVH health indicators. Due to the strong accessibility of health behavior assessment in patients’ daily life, the CVH status based on LE8 may reduce the risk of death and improve their quality of life.

There has been controversy in previous large cohort studies about whether lifestyle interventions are effective in reducing the all-cause mortality in diabetic patients. We applied a uniform CVH quantification criterion based on LE8 and showed that high CVH status was significantly associated with reduced all-cause mortality in prediabetic patients. Effective health interventions are important for improving the survival prognosis and reducing the medical burden of prediabetes, the results of the Da Qing Diabetes Prevention Outcomes Study (DQDPOS) support our view.

Previous studies have shown that quantified CVH has a protective effect on diabetes incidence, quality of life and mortality^20, 30–33^. In a follow-up cohort of 309,789 participants, Sun et al. found that higher CVH status was associated with reduced premature mortality among T2DM patients (HR=0.42,95%CI=0.39-0.45)^21^. Based on the guidelines from American Association of Clinical Endocrinology and considering the value of early life interventions, we further studied patients with prediabetes and found a significant protective effect. In a meta-analysis of 193,126 participants, Geidl et al. found that a 1-point improvement in CVH status based on the LS7 score was associated with an average 11% reduction in all-cause mortality (RR=0.89,95%CI=0.86-0.93) observing significant dose-response characteristics^34^. However, LS7 quantifies CVH by simply adding categorical scores of seven factors, which may result in insufficient statistical power for dose-response associations^20^. Therefore, we used LE8 from the mean score of the hundred-mark scale for the eight health factors for the analysis and found significant dose-response associations (P-overall < 0.0001,P-nonliner = 0.7989). Every 10-point improvement in LE8 score was associated with a 30% reduction in prediabetic patients’ all-cause mortality, which means that even a slight improvement in CVH status may have a significant effect on the prevention of mortality in prediabetic patients. Given the accessibility of the various assessments of the LE8 in the daily life of patients, we focused on the health behavior part of CVH. The results showed that health behaviors could have more significant improvement on the survival prognosis of patients. However, after adjustments, the health indicator scores have no significant protective effect, which may be due to a more fundamental effect of health behaviors. Our results may suggest that physicians should strengthen health behavior education for patients with prediabetes. Marteau et al. highlighted that promoting diet, physical activity and other behaviors into the healthiest state, remains a major challenge for cardiometabolic disease prevention efforts, which further supports our view^35^.

Among CVH health behaviors, we noticed that increased physical activity was the top protective factor in reducing the risk of all-cause mortality. The protective association of physical activity has been widely demonstrated^30^, and our analysis validates the recent scoring method of physical activity in LE8 in the prediabetic population. We also suggest that nicotine exposure was the second most significant contributor to prediabetic patient’s mortality. Cohort study data show that smoking cessation is associated with reduced risk for all-cause mortality^36^. Our findings confirmed this association and emphasize the protective significance of long-term maintenance of smoking cessation status in prediabetic patients with a history of smoking, based on the LE8-based scoring method for the duration of smoking cessation. Previous evidence has shown that unhealthy sleep duration (<7 or >8 hours) is associated with increased all-cause mortality^37^. Our results showed that the risk of all-cause mortality decreased significantly with the increase of sleep index score.

Interestingly, a significant positive association was shown between BMI score and blood lipid score greater than 50 and all-cause mortality in prediabete patients. This may be related to the obesity paradox effect, where obesity appears as a risk factor in disease incidence but has a protective effect in patients with severe disease^25, 40–42^. Similarly, in a meta-analysis of 161,984 participants, Liu et al. observed the protective effect of overweight in patients with type 2 diabetes^43^. However, an opposite result was observed in a follow-up study with an average of 15.8 years, and Tobias et al. suggested that the obesity paradox was due to insufficient follow-up time^44^. Our findings may provide new evidence in this controversial area, and further studies are needed to confirm possible interactions in the prediabetic population. Our results noticed that the relationship between dietary health score ≥ 50 and all-cause mortality was not significant. Because there were few participants with a score of 80 or higher in each category, we used a cutoff of 50 or higher to improve statistical power. This cutoff may not have met the health requirements for improved outcomes in patients with prediabetes, given that diet quality is a major determinant of the risk of diabetes^38, 39^. In a cohort study, also based on a US population, Fretts et al. observed a similar phenomenon^33^. A similar reason may explain the nonsignificant results for blood-pressure scores.

To the best of our knowledge, this is the first study to examine the association between quantified CVH and all-cause mortality in prediabetic patients. The strength of this study is that it considers both the need to prevent prediabetes and the need to mitigate the impact of CVD comorbidities on prediabetic patients. In addition, we designed the study with a relatively large sample size and long-term follow-up, used the new LE8 score recommended by the AHA to accurately quantify CVH status, and performed covariate adjustment and sensitivity analysis during the data analysis, which makes our results more reliable.

However, there are still several potential limitations worth considering. First, this study was an observational cohort study, so causality should be interpreted with caution. Secondly, the four CVH health behaviors (diet, physical activity, nicotine exposure, and sleep) in the LE8 score were all self-reported, which may lead to recall bias. More objective monitoring would be beneficial to improve the accuracy of these four health behaviors. Third, we obtained the CVH score only once at baseline, which may change during long-term follow-up. Finally, our study population included only U.S. adults, and generalization of the results to other populations should be cautious.

## **V.** Conclusion

We suggest that a high CVH status based on LE8 quantification, especially CVH health behaviors in prediabetic adults has a significant effect in preventing mortality outcomes in prediabetic adults. There was a linear dose-response association between increased LE8 and reduced all-cause mortality. Our findings highlight the importance of focusing on CVH improvement in prediabetic patients, especially on more accessible health behaviors, as even small improvements can yield substantial reductions in the risk of all-cause mortality.

## Data Availability

All data produced are available online at NCHS:https://www.cdc.gov/nchs/nhanes/index.htm

NCHS:https://www.cdc.gov/nchs/nhanes/index.htm

## Acknowledgement

This research has been conducted using the NHANES resources. We are grateful to the participants and staff of the NHANES for their valuable contributions.

## Funding

This work was supported by research grants from the National Natural Science Foundation of China (Y.J., 82370818, Z.Z., 82270862); Natural Science Foundation of Guangdong Province(Y.J.,2024A1515012744); the Guangzhou Science and Technology Project (Y.J.,2024A04J5098); and the National Undergraduate Training Program for Innovation and Entrepreneurship, Southern Medical University (Z.Z., 202312121031, Y.J., S202312121167).

## Author contributions

A.C., Q.H., Y.W., Z.Z., and Y.J. designed this study, A.C., Y.W. and Q.H. performed the data extraction and statistical analysis. Y.W., Q.H., J.C., X.M., L.H., G.W., and Z.W. validated the statistical analysis. A.C. and Q.H. wrote the original draft of the manuscript, which was revised by Y.W., Z.Z., and Y.J. All authors made great contributions to the manuscript and approved it for submission. Z.Z. and Y.J. are the guarantors of this work and, as much, had full access to all the data in the study and take responsibility for the integrity of the data and accuracy of the data analysis.

## Conflict of interest

All authors declare no competing interests.

## Notes

### Competing Interest Statement

The authors have declared no competing interest.

### Author Declarations

The study used (or will use) ONLY openly available human data that were originally located at:NCHS:https://www.cdc.gov/nchs/nhanes/index.htm

## References

1 Tabák AG, Herder C, Rathmann W, Brunner EJ, Kivimäki M. Prediabetes: a high-risk state for diabetes development. The Lancet. 2012;379(9833):2279-2290.

2 Cai X, Zhang Y, Li M, Wu JH, Mai L, Li J, Yang Y, Hu Y, Huang Y. Association between prediabetes and risk of all cause mortality and cardiovascular disease: updated meta-analysis. BMJ. 2020:m2297.

3 Kim SH. Reframing prediabetes: A call for better risk stratification and intervention. J INTERN MED. 2024;295(6):735–747.

4. 3. Prevention or Delay of Type 2 Diabetes:Standards of Medical Care in Diabetes — 2020. DIABETES CARE. 2020;43(Supplement_1):S32-S36.

5 Cosentino F, Grant PJ, Aboyans V, Bailey CJ, Ceriello A, Delgado V, Federici M, Filippatos G, Grobbee DE, Hansen TB, Huikuri HV, Johansson I, Jüni P, Lettino M, Marx N, Mellbin LG, Östgren CJ, Rocca B, Roffi M, Sattar N, Seferović PM, Sousa-Uva M, Valensi P, Wheeler DC, Piepoli MF, Birkeland KI, Adamopoulos S, Ajjan R, Avogaro A, Baigent C, Brodmann M, Bueno H, Ceconi C, Chioncel O, Coats A, Collet J, Collins P, Cosyns B, Di Mario C, Fisher M, Fitzsimons D, Halvorsen S, Hansen D, Hoes A, Holt RIG, Home P, Katus HA, Khunti K, Komajda M, Lambrinou E, Landmesser U, Lewis BS, Linde C, Lorusso R, Mach F, Mueller C, Neumann F, Persson F, Petersen SE, Petronio AS, Richter DJ, Rosano GMC, Rossing P, Rydén L, Shlyakhto E, Simpson IA, Touyz RM, Wijns W, Wilhelm M, Williams B, Aboyans V, Bailey CJ, Ceriello A, Delgado V, Federici M, Filippatos G, Grobbee DE, Hansen TB, Huikuri HV, Johansson I, Jüni P, Lettino M, Marx N, Mellbin LG, Östgren CJ, Rocca B, Roffi M, Sattar N, Seferović PM, Sousa-Uva M, Valensi P, Wheeler DC, Windecker S, Aboyans V, Baigent C, Collet J, Dean V, Delgado V, Fitzsimons D, Gale CP, Grobbee DE, Halvorsen S, Hindricks G, Iung B, Jüni P, Katus HA, Landmesser U, Leclercq C, Lettino M, Lewis BS, Merkely B, Mueller C, Petersen SE, Petronio AS, Richter DJ, Roffi M, Shlyakhto E, Simpson IA, Sousa-Uva M, Touyz RM, Zelveian PH, Scherr D, Jahangirov T, Lazareva I, Shivalkar B, Naser N, Gruev I, Milicic D, Petrou PM, Linhart A, Hildebrandt P, Hasan-Ali H, Marandi T, Lehto S, Mansourati J, Kurashvili R, Siasos G, Lengyel C, Thrainsdottir IS, Aronson D, Di Lenarda A, Raissova A, Ibrahimi P, Abilova S, Trusinskis K, Saade G, Benlamin H, Petrulioniene Z, Banu C, Magri CJ, David L, Boskovic A, Alami M, Liem AH, Bosevski M, Svingen GFT, Janion M, Gavina C, Vinereanu D, Nedogoda S, Mancini T, Ilic MD, Fabryova L, Fras Z, Jiménez-Navarro MF, Norhammar A, Lehmann R, Mourali MS, Ural D, Nesukay E, Chowdhury TA. 2019 ESC Guidelines on diabetes, pre-diabetes, and cardiovascular diseases developed in collaboration with the EASD. EUR HEART J. 2020;41(2):255-323.

6 Huang Y, Cai X, Mai W, Li M, Hu Y. Association between prediabetes and risk of cardiovascular disease and all cause mortality: systematic review and meta-analysis. BMJ. 2016:i5953.

7. Diagnosis and classification of diabetes mellitus. DIABETES CARE. 2014;37 Suppl 1:S81-S90.

8 Gong Q, Zhang P, Wang J, Ma J, An Y, Chen Y, Zhang B, Feng X, Li H, Chen X, Cheng YJ, Gregg EW, Hu Y, Bennett PH, Li G, Qian X, Zhang L, Hui Y, He S, Wang X, Thompson TJ, Gerzoff RB, Liu P, Jiang Y, Hu Z, Wang J, Jiang X, Zhang J, Xi R, Pang C, Li C, Hu X, Yang W, An Z, Sun X, Chen C, Gang Y, Liu J, Xiao J, Cao H, Zheng H, Zhang H, Li H, Hong J, Liu X, Zhao F, Wang W, Chen B, Howard BV, Engelgau MM, Roglic G. Morbidity and mortality after lifestyle intervention for people with impaired glucose tolerance: 30-year results of the Da Qing Diabetes Prevention Outcome Study. The Lancet Diabetes & Endocrinology. 2019;7(6):452–461.

9 Lee CG, Heckman-Stoddard B, Dabelea D, Gadde KM, Ehrmann D, Ford L, Prorok P, Boyko EJ, Pi-Sunyer X, Wallia A, Knowler WC, Crandall JP, Temprosa M. Effect of Metformin and Lifestyle Interventions on Mortality in the Diabetes Prevention Program and Diabetes Prevention Program Outcomes Study. DIABETES CARE. 2021;44(12):2775–2782.

10. Effects of Intensive Lifestyle Intervention on All-Cause Mortality in Older Adults With Type 2 Diabetes and Overweight/Obesity: Results From the Look AHEAD Study. DIABETES CARE. 2022;45(5):1252-1259.

11 Zhang Y, Pan X, Lu Q, Wang Y, Geng T, Zhou Y, Liao LM, Tu Z, Chen J, Xia P, Wang Y, Wan Z, Guo K, Yang K, Yang H, Chen S, Wang G, Han X, Wang Y, Yu D, He M, Zhang X, Liu L, Wu T, Wu S, Liu G, Pan A. Association of Combined Healthy Lifestyles With Cardiovascular Disease and Mortality of Patients With Diabetes: An International Multicohort Study. MAYO CLIN PROC. 2023;98(1):60–74.

12. Effect of Metformin and Lifestyle Interventions on Mortality in the Diabetes Prevention Program and Diabetes Prevention Program Outcomes Study.

13 Pan XR, Li GW, Hu YH, Wang JX, Yang WY, An ZX, Hu ZX, Lin J, Xiao JZ, Cao HB, Liu PA, Jiang XG, Jiang YY, Wang JP, Zheng H, Zhang H, Bennett PH, Howard BV. Effects of diet and exercise in preventing NIDDM in people with impaired glucose tolerance. The Da Qing IGT and Diabetes Study. DIABETES CARE. 1997;20(4):537–544.

14 He Y, Wang Z, Zhang H, Lai X, Liu M, Yang L, Zheng Y, He M, Kong W, Zhang X. Polygenic Risk Score Modifies the Association of HbA1c With Hearing Loss in Middle-Aged and Older Chinese: The Dongfeng-Tongji Cohort. DIABETES CARE. 2024.

15 Lloyd-Jones DM, Allen NB, Anderson CAM, Black T, Brewer LC, Foraker RE, Grandner MA, Lavretsky H, Perak AM, Sharma G, Rosamond W. Life’ s Essential 8: Updating and Enhancing the American Heart Association’s Construct of Cardiovascular Health: A Presidential Advisory From the American Heart Association. CIRCULATION. 2022;146(5).

16 Tang R, Wang X, Li X, Ma H, Liang Z, Heianza Y, Qi L. Adherence to Life’s Essential 8 and incident chronic kidney disease: a prospective study of 147,988 UK Biobank participants. The American Journal of Clinical Nutrition. 2023;118(4):804–811.

17 Kulkarni AV, Anders M, Nazal L, Ridruejo E, Efe C. Cases of severe acute liver injury following inactivated SARS-CoV-2 vaccination. J HEPATOL. 2023;78(2):e60–e61.

18 Wu S, Wu Z, Yu D, Chen S, Wang A, Wang A, Gao X. Life’s Essential 8 and Risk of Stroke: A Prospective Community-Based Study. STROKE. 2023;54(9):2369–2379.

19 Janssen I, Powell LH, Dugan SA, Derby CA, Kravitz HM. Cardiovascular Health, Race, and Decline in Cognitive Function in Midlife Women: The Study of Women’s Health Across the Nation. J AM HEART ASSOC. 2024;13(9).

20 Sun J, Li Y, Zhao M, Yu X, Zhang C, Magnussen CG, Xi B. Association of the American Heart Association’s new “Life’s Essential 8” with all-cause and cardiovascular disease-specific mortality: prospective cohort study. BMC MED. 2023;21(1).

21 Sun Y, Yu Y, Zhang K, Yu B, Yu Y, Wang Y, Wang B, Tan X, Wang Y, Lu Y, Wang N. Association between Life’s Essential 8 score and risk of premature mortality in people with and without type 2 diabetes: A prospective cohort study. Diabetes/Metabolism Research and Reviews. 2023;39(5).

22 Lloyd-Jones DM, Ning H, Labarthe D, Brewer L, Sharma G, Rosamond W, Foraker RE, Black T, Grandner MA, Allen NB, Anderson C, Lavretsky H, Perak AM. Status of Cardiovascular Health in US Adults and Children Using the American Heart Association’s New “Life’s Essential 8” Metrics: Prevalence Estimates From the National Health and Nutrition Examination Survey (NHANES), 2013 Through 2018. CIRCULATION. 2022;146(11):822-835.

23 Boyer TM, Avula V, Minhas AS, Vaught AJ, Sharma G, Gemmill A. Psychosocial Stressors as a Determinant of Maternal Cardiovascular Health During Pregnancy. The American Journal of Cardiology. 2023;201:302–307.

24 Zhang H, Zhang L, Li J, Xiang H, Liu Y, Gao C, Sun X. The influence of Life’s Essential 8 on the link between socioeconomic status and depression in adults: a mediation analysis. BMC PSYCHIATRY. 2024;24(1).

25 Zheng J, Hu Y, Xu H, Lei Y, Zhang J, Zheng Q, Li L, Tu W, Chen R, Guo Q, Zang X, You Q, Xu Z, Zhou Q, Wu X. Normal-weight visceral obesity promotes a higher 10-year atherosclerotic cardiovascular disease risk in patients with type 2 diabetes mellitus – a multicenter study in China. CARDIOVASC DIABETOL. 2023;22(1).

26 Ueno K, Kaneko H, Okada A, Suzuki Y, Matsuoka S, Fujiu K, Michihata N, Jo T, Takeda N, Morita H, Kamiya K, Ako J, Node K, Yasunaga H, Komuro I. Association of four health behaviors in Life’s Essential 8 with the incidence of hypertension and diabetes mellitus. PREV MED. 2023;175:107685.

27 Zhang Q, Xiao S, Jiao X, Shen Y. The triglyceride-glucose index is a predictor for cardiovascular and all-cause mortality in CVD patients with diabetes or pre-diabetes: evidence from NHANES 2001– 2018. CARDIOVASC DIABETOL. 2023;22(1).

28 Zou X, Zhou X, Zhu Z, Ji L. Novel subgroups of patients with adult-onset diabetes in Chinese and US populations. Lancet Diabetes Endocrinol. 2019;7(1):9–11.

29 Shams-White MM, Pannucci TE, Lerman JL, Herrick KA, Zimmer M, Meyers MK, Stoody EE, Reedy J. Healthy Eating Index-2020: Review and Update Process to Reflect the Dietary Guidelines for Americans,2020-2025. J ACAD NUTR DIET. 2023;123(9):1280-1288.

30 Yang Q, Cogswell ME, Flanders WD, Hong Y, Zhang Z, Loustalot F, Gillespie C, Merritt R, Hu FB. Trends in Cardiovascular Health Metrics and Associations With All-Cause and CVD Mortality Among US Adults. JAMA. 2012;307(12):1273.

31 Perak AM, Ning H, Khan SS, Bundy JD, Allen NB, Lewis CE, Jacobs DR, Van Horn LV, Lloyd-Jones DM. Associations of Late Adolescent or Young Adult Cardiovascular Health With Premature Cardiovascular Disease and Mortality. J AM COLL CARDIOL. 2020;76(23):2695–2707.

32 Xanthakis V, Enserro DM, Murabito JM, Polak JF, Wollert KC, Januzzi JL, Wang TJ, Tofler G, Vasan RS. Ideal Cardiovascular Health. CIRCULATION. 2014;130(19):1676–1683.

33 FRETTS AM, HOWARD BV, MCKNIGHT B, DUNCAN GE, BERESFORD SAA, METE M, ZHANG Y, SISCOVICK DS. Life’s Simple 7 and Incidence of Diabetes Among American Indians: The Strong Heart Family Study. DIABETES CARE. 2014;37(8):2240–2245.

34 Geidl W, Schlesinger S, Mino E, Miranda L, Pfeifer K. Dose – response relationship between physical activity and mortality in adults with noncommunicable diseases: a systematic review and meta-analysis of prospective observational studies. INT J BEHAV NUTR PHY. 2020;17(1).

35 Marteau TM, Hollands GJ, Fletcher PC. Changing Human Behavior to Prevent Disease: The Importance of Targeting Automatic Processes. SCIENCE. 2012;337(6101):1492-1495.

36 Han L, You D, Ma W, Astell-Burt T, Feng X, Duan S, Qi L. National Trends in American Heart Association Revised Life’s Simple 7 Metrics Associated With Risk of Mortality Among US Adults. JAMA Network Open. 2019;2(10):e1913131.

37 Krittanawong C, Tunhasiriwet A, Wang Z, Zhang H, Farrell AM, Chirapongsathorn S, Sun T, Kitai T, Argulian E. Association between short and long sleep durations and cardiovascular outcomes: a systematic review and meta-analysis. European Heart Journal: Acute Cardiovascular Care. 2019;8(8):762–770.

38 Schwingshackl L, Hoffmann G, Lampousi A, Knüppel S, Iqbal K, Schwedhelm C, Bechthold A, Schlesinger S, Boeing H. Food groups and risk of type 2 diabetes mellitus: a systematic review and meta-analysis of prospective studies. EUR J EPIDEMIOL. 2017;32(5):363–375.

39 Wang B, Fu Y, Tan X, Wang N, Qi L, Lu Y. Assessing the impact of type 2 diabetes on mortality and life expectancy according to the number of risk factor targets achieved: an observational study. BMC MED. 2024;22(1).

40 Ottosson F, Smith E, Ericson U, Brunkwall L, Orho-Melander M, Di Somma S, Antonini P, Nilsson PM, Fernandez C, Melander O. Metabolome-Defined Obesity and the Risk of Future Type 2 Diabetes and Mortality. DIABETES CARE. 2022;45(5):1260–1267.

41 Al-Chalabi S, Chinnadurai R, Kalra PA, Sinha S. Mortality and Renal Outcomes Are Impacted by Obesity in Cardiorenal Metabolic Disease but Not in People with Concomitant Diabetes Mellitus. CARDIORENAL MED. 2024;14(1):23–33.

42 Nie W, Zhang Y, Jee SH, Jung KJ, Li B, Xiu Q. Obesity survival paradox in pneumonia: a meta-analysis. BMC MED. 2014;12:61.

43 Liu X, Liu Y, Zhan J, He Q. Overweight, obesity and risk of all-cause and cardiovascular mortality in patients with type 2 diabetes mellitus: a dose – response meta-analysis of prospective cohort studies. EUR J EPIDEMIOL. 2015;30(1):35–45.

44 Tobias DK, Pan A, Jackson CL, O’Reilly EJ, Ding EL, Willett WC, Manson JE, Hu FB. Body-Mass Index and Mortality among Adults with Incident Type 2 Diabetes. NEW ENGL J MED. 2014;370(3):233–244.

