## Supplementary Table 1 for "Association of quantified cardiovascular health with all-cause mortality in prediabetic patients"

**Table 1-Baseline characteristics of the study population classified by CVH status.**

| **Characteristics** | **Overall** | **CVH status (LE8 score)** | | | **p** |
| --- | --- | --- | --- | --- | --- |
|  |  | **High(80-100)** | **Moderate(50-79)** | **Low(0-49)** |  |
| **Weighted N** | 226 million | 7.0 (15.8 million) | 76.7 (173.3 million) | 13.6 (36.9 million) |  |
| **Age , years** | 52.86 (15.87) | 44.73 (17.38) | 53.24 (15.93) | 54.58 (13.75) | <0.001 |
| **Sex** |  |  |  |  | 0.008 |
| Male | 51.6 (116.6 million ) | 57.4 (9.1 million ) | 52.3 (90.7 million ) | 45.7 (16.8 million ) |  |
| Female | 48.4 (109.4 million ) | 42.6 (6.7 million ) | 47.7 (82.6 million ) | 54.3 (20 million ) |  |
| **Ethnicity** |  |  |  |  | 0.01 |
| Black | 15.1 (34 million ) | 11.0 (1.7 million ) | 14.7 (25.5 million ) | 18.3 (6.8 million ) |  |
| Other | 20.2 (45.6 million ) | 25.7 (4.1 million ) | 20.1 (34.9 million ) | 18.1 (6.7 million ) |  |
| White | 64.7 (146.3 million ) | 63.3 (10 million ) | 65.1 (112.9 million ) | 63.6 (23.4 million ) |  |
| **PIR*** | 2.97(1.64) | 3.30(1.59 ) | 3.05(1.63) | 2.34(1.58) | <0.001 |
| **Drinker*** | 59.4 (134.3 million ) | 53.6 (8.5 million ) | 59.0 (102.3 million ) | 63.9 (23.6 million ) | 0.049 |
| **Smoker*** | 48.6 (109.7 million ) | 21.7 (3.4 million ) | 45.3 (78.5 million ) | 75.4 (27.8 million ) | <0.001 |
| **BMI** | 30.14 (6.83) | 24.80 (3.75) | 29.72 (6.37) | 34.40 (7.63) | <0.001 |
| **Sleep duration, hours** | 6.77 (1.66) | 7.20 (0.91) | 6.82 (1.21) | 6.35 (3.06) | <0.001 |
| **Creatinine, mmol/L** | 0.89 (0.38) | 0.87 (0.19) | 0.90 (0.41) | 0.87 (0.26) | 0.151 |
| **Glucose, mmol/L** | 102.19 (20.34) | 95.73 (10.04) | 101.58 (18.67) | 107.81 (28.36) | <0.001 |
| **TC, mmol/L** | 196.91 (41.58) | 175.14 (29.26) | 194.84 (40.58) | 216.00 (43.65) | <0.001 |
| **SBP,mmHg** | 124.06 (17.00) | 114.90 (12.86) | 123.56 (16.29) | 130.34 (19.41) | <0.001 |
| **DBP,mmHg** | 69.95 (12.56) | 67.58 (8.87) | 69.57 (12.37) | 72.73 (14.26) | <0.001 |
| **CVD** | 9.3 (21 million ) | 7.2 (1.1 million ) | 8.7 (15.1 million ) | 13.0 (4.8 million ) | 0.006 |
| **Diabetes** | 17.8 (40.3 million ) | 0.7 (0.1 million ) | 16.1 (27.9 million ) | 33.3 (12.3 million ) | <0.001 |
| **Hypertension** | 38.1 (86.1 million ) | 14.2 (2.3 million ) | 37.6 (65.2 million ) | 50.4 (18.6 million ) | <0.001 |
| **Anemia** | 1.8 (4.1 million ) | 0.6 (0.1 million ) | 1.8 (3.1 million ) | 2.5 (0.9 million ) | <0.001 |
| **LE8** | 62.17 (12.03) | 83.77 (3.16) | 64.12 (8.02) | 43.76 (4.91) | <0.001 |

*Note*：Data are %（weighted N） for categorical measures or mean (SD) for continuous measures. *PIR :the ratio of family income to poverty; Smoker:smoked at least 100 cigarettes in lifetime; Drinker: had at least 12 alcohol drinkers in lifetime; TC:total cholesterol.

**Table 2-Hazard ratios of cardiovascular health status to all-cause mortality in prediabetes.**

|  |  | **Model1** |  |  | **Model2** |  |  | **Model3** |  |
| --- | --- | --- | --- | --- | --- | --- | --- | --- | --- |
| **CVH status(LE8 score)** | **N** | **HR(95%CI)** | **P-value** |  | **HR(95%CI)** | **P-value** |  | **HR(95%CI)** | **P-value** |
| High(80-100) | 199 | Reference |  |  | Reference |  |  | Reference |  |
| Moderate(50-79) | 3943 | 4.32(2.10,8.91) | <0.001 |  | 2.68(1.34，5.36) | 0.005 |  | 2.55(1.23，5.31) | 0.012 |
| Low(0-49) | 1202 | 8.39(4.03,17.48) | <0.001 |  | 4.75(2.27，9.93) | <0.001 |  | 3.92(1.70，9.02) | 0.001 |

*Note：*Model1：No adjustment. Model2：Adjusted for sex, age, race/ethnicity, and income. Model3：Adjusted for sex, age, race/ethnicity, income,alcohol consumption, smoking, BMI, total cholesterol, diabetes, anemia, hypertension, and cardiovascular disease.

**Supplementary Table 1:** Methods for evaluating each individual cardiovascular health metric.

| **Domain** | **CVH metric** | **Method of measurement** | **Quantification of CVH metric:adult（≥20 years of age）** | |
| --- | --- | --- | --- | --- |
| **Health behaviors** | **Diet** |  | Points | Metrics: Healthy Eating Index-2020 |
|  |  | Healthy Eating Index-2020 was assessed using 2 interviewer-administered 24-hour dietary recalls | 100 | 95th percentile (top/ideal diet) |
|  |  |  | 80 | 75th–94th percentile |
|  |  |  | 50 | 50th–74th percentile |
|  |  |  | 25 | 25th–49th percentile |
|  |  |  | 0 | 1st–24th percentile (bottom/ least ideal quartile) |
|  | **Physical activity** |  | Points | Metrics: minutes of moderate to vigorous physical activity per week |
|  |  | Self-reported minutes of moderate or vigorous physical activity per week | 100 | ≥150 minutes |
|  |  |  | 90 | 120-149 minutes |
|  |  |  | 80 | 90-119 minutes |
|  |  |  | 60 | 60-89 minutes |
|  |  |  | 40 | 30-59 minutes |
|  |  |  | 20 | 1-29 minutes |
|  |  |  | 0 | 0 minutes |
|  | **Tobacco/nicotine exposure** |  | Points | Metrics: Combustible tobacco use or inhaled NDS use; or secondhand smoke exposure |
|  |  | Self-reported use of cigarettes or inhaled nicotine-delivery system (NDS), or secondhand smoke exposure | 100 | Never smoker |
|  |  |  | 75 | Former smoker, quit ≥ 5 years |
|  |  |  | 50 | Former smoker, quit 1–<5 years |
|  |  |  | 25 | Former smoker, quit <1 years, or currently using inhaled NDS |
|  |  |  | 0 | Current smoker |
|  |  |  | Subtract 20 points (unless score is 0) for living with active indoor smoker in home | |
|  | **Sleep health** |  | Points | Metrics: Average hours of sleep per night |
|  |  | Self-reported average  hours of sleep per night | 100 | 7-9 hours |
|  |  |  | 90 | 9-<10 hours |
|  |  |  | 70 | 6–<7 hours |
|  |  |  | 40 | 5–<6 or ≥10 hours |
|  |  |  | 20 | 4–<5 hours |
|  |  |  | 0 | < 4 hours |
| **Health indicators** | **Body mass index** |  | Points | Metrics: BMI |
|  |  | Body mass index (BMI) was calculated as the weight (kilograms) divided by the square of the height (meters squared) from standardized height and weight measurements. | 100 | <25 kg/m^2^ |
|  |  |  | 70 | 25.0-29.9 kg/m^2^ |
|  |  |  | 30 | 30.0-34.9 kg/m^2^ |
|  |  |  | 15 | 35.0-39.9 kg/m^2^ |
|  |  |  | 0 | ≥ 40.0 kg/m^2^ |
|  | **Blood lipid** |  | Points | Metrics: Non-HDL cholesterol |
|  |  | Non-HDL cholesterol was calculated by total cholesterol minus HDL cholesterol using fasting or nonfasting blood sample | 100 | <130 mg/dL |
|  |  |  | 60 | 130-159 mg/dL |
|  |  |  | 40 | 160-189 mg/dL |
|  |  |  | 20 | 190-219 mg/dL |
|  |  |  | 0 | ≥ 220 mg/dL |
|  |  |  | If drug-treated level, subtract 20 points | |
|  | **Blood glucose** |  | Points | Metrics: FBG or HbA1c |
|  |  | hemoglobin A1c (HbA1c) was measured by high-performance liquid chromatography methods.  Fasting blood glucose (FBG) was measured by standard methods. | 100 | No history of diabetes and  FBG <100 mg/dL (or HbA1c <5.7 %) |
|  |  |  | 60 | No diabetes and FBG 100–125 100 mg/dL (or HbA1c 5.7–6.4%) |
|  |  |  | 40 | Diabetes with HbA1c <7.0 % |
|  |  |  | 30 | Diabetes with HbA1c 7.0–7.9 % |
|  |  |  | 20 | Diabetes with HbA1c 8.0–8.9 % |
|  |  |  | 10 | Diabetes with HbA1c 9.0–9.9 % |
|  |  |  | 0 | Diabetes with HbA1c ≥10.0 % |
|  | **Blood pressure** |  | Points | Metrics: Systolic and diastolic BPs |
|  |  | The average of all available blood pressure (BP) measurements was used to calculate systolic and diastolic BP. BPs were measured in mobile examination centers with standard protocols. | 100 | <120/<80 mm Hg (optimal) |
|  |  |  | 75 | 120-129/<80 mm Hg (elevated) |
|  |  |  | 50 | 130-139 or 80-89 mm Hg (stage 1 hypertension) |
|  |  |  | 25 | 140-159 or 90-99 mm Hg |
|  |  |  | 0 | ≥ 160 or ≥ 100 mm Hg |
|  |  |  | Subtract 20 points (unless score is 0) if treated level | |

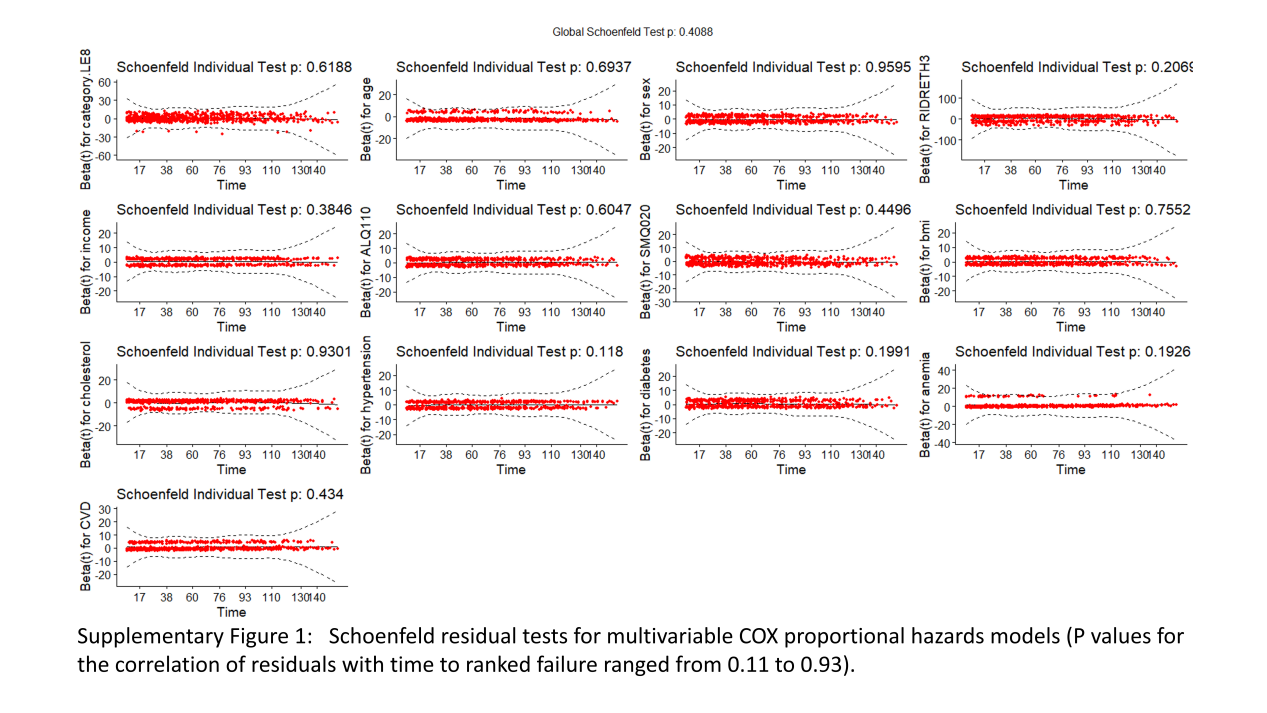

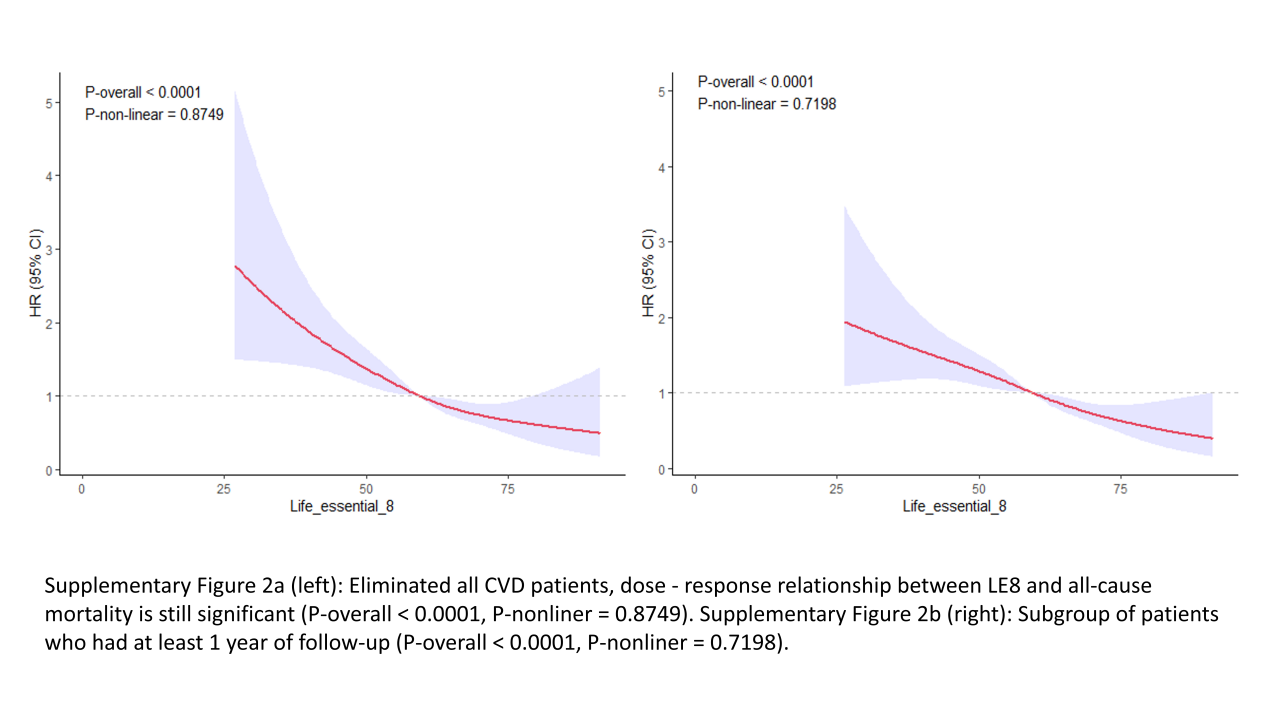
